# Risk of death following SARS-CoV-2 infection or COVID-19 vaccination in young people in England: a self-controlled case series study

**DOI:** 10.1101/2022.03.22.22272775

**Authors:** Vahé Nafilyan, Charlotte Bermingham, Isobel L. Ward, Jasper Morgan, Francesco Zaccardi, Kamlesh Khunti, Julie Stanborough, Amitava Banerjee

## Abstract

**Objectives:** To assess whether there is a change in the incidence of cardiac and all-cause death in young people following COVID-19 vaccination or SARS-CoV-2 infection in unvaccinated individuals.

**Design:** Self-controlled case series.

**Setting:** National, linked electronic health record data in England.

**Study population:** Individuals aged 12-29 who had received at least one dose of COVID-19 vaccination and died between 8 December 2020 and 2 February 2022 and registered by 16 February 2022 within 12 weeks of COVID-19 vaccination; Individuals aged 12-29 who died within 12 weeks of testing positive for SARS-CoV-2.

**Main outcome measures:** Cardiac and all-cause deaths occurring within 12 weeks of vaccination or SARS-CoV-2 infection.

**Results:** Compared to the baseline period, there was no evidence of a change in the incidence of cardiac death in the six weeks after vaccination, whether for each of weeks 1 to 6 or the whole six-week period. There was a decrease in the risk of all-cause death in the first week after vaccination and no change in each of weeks 2 to 6 after vaccination or whole six-week period after vaccination. Subgroup analyses by sex, age, vaccine type, and last dose also showed no change in the risk of death in the first six weeks after vaccination. There was a large increase in the incidence of cardiac and all-cause death in the overall risk period after SARS-CoV-2 infection among the unvaccinated.

**Conclusion:** There is no evidence of an association between COVID-19 vaccination and an increased risk of death in young people. By contrast, SARS-CoV-2 infection was associated with substantially higher risk of cardiac related death and all-cause death.

**What is already known on this topic:** Several studies have highlighted the association between COVID-19 vaccination and the risk of myocarditis, myopericarditis, and other cardiac problems, especially in young people, but associated risk of mortality is unclear. Since younger people have lower risk of COVID-19 hospitalisation and mortality, the mortality risk associated with vaccination is potentially more important to them in balancing the risk and benefit of vaccination.

**What this study adds:** Although there is a risk of myocarditis or myopericarditis with COVID-19, there is no evidence of increased risk of cardiac or all-cause mortality following COVID-19 vaccination in young people aged 12 to 29. Given the increased risk of mortality following SARS-CoV-2 infection in this group, the risk-benefit analysis favours COVID-19 vaccination for this age group.

## Introduction

On the 8 December 2020 the UK began administering vaccines against COVID-19 according to the priority groups determined by the Joint Committee on Vaccination and Immunisation (JCVI). The vaccines administered have been shown to have high efficacy in trials and effectiveness in routine care against death and hospitalisation due to COVID-19 [1, 2, 3]. However, it is also important to consider their safety, which is difficult to assess in randomised clinical trials as these are not powered to detect rare adverse events [4, 5].

There have been rare cases of serious side effects reported with the COVID-19 vaccine. Previous studies have shown an increase in the risk of myocarditis and myopericarditis associated with mRNA vaccines including BNT162b2 (Pfizer-BioNTech) and mRNA-1273 (Moderna) [1, 6] and an increased risk of thrombotic events after the ChAdOx1 nCoV-19 vaccine (Oxford-AstraZeneca) [7]. However, the absolute risk was low and should be balanced against the risk of the event occurring after SARS-CoV-2 infection [8, 9]. The balance of risk and benefit is particularly important to determine for younger people, due to the lower risk of COVID-19 hospitalisation and death in this age group [10].

Comparisons of the risk of death in vaccinated and unvaccinated young people are subject to confounding particularly due to the prioritisation, among younger people, of those with underlying health conditions. In England, 16-64 year olds with underlying health conditions became eligible for vaccination on 15 February 2021 whilst the general eligibility for younger ages began on 18 June 2021 for adults aged 18 years and over, followed by 16-17 year olds on 4 August 2021, 12-15 year olds on 13 September, and 5-11 year olds on 15 February 2022.

We therefore used a self-controlled case study design, where each participant acts as their own control, to compare the risk of death soon after vaccination (‘risk period’) to a baseline period after the risk period [11]. For comparison purposes, we also assessed the relative incidence of death following SARS-CoV-2 infection in unvaccinated individuals.

## Methods

### Data sources

We linked death registration data from the Office for National Statistics to data on COVID-19 vaccination from the National Immunisation Management Service (NIMS) and an extract from NHS point of care data provided by NHS-Digital. The NIMS data includes most COVID-19 vaccinations administered in England since 8 December 2020. However, in some rare cases, the vaccination records of people who died shortly after vaccination may not be recorded in NIMS. This would happen if the death was recorded on the Personal Demographics Service (PDS) before the vaccination records were sent to NIMS. Therefore, we supplemented the vaccination records from NIMS using a special extract of 2,044 people who died after vaccination but whose records for the last vaccination received were not sent to NIMS, 2 of whom were aged 12-29 and had a linked death record. To assess the relative incidence of death following SARS-CoV-2 infection, we also used national testing data from pillar 1 (tests in hospitals) and pillar 2 (tests in the community).

The linkage was conducted using NHS number, which was available for 99.96% of NIMS records, 99.6% of deaths and 100% of the extract from NHS-Digital.

The data covers people resident in England and included deaths that occurred between 8 December 2020 and 2 February 2022 and were registered by 16 February 2022,vaccinations that were recorded up to 16 February 2022 and infections recorded up to 30 December 2021.

### Study population

The study population included all people who died within 12 weeks of having received their last dose (first, second, or third dose or booster) of a COVID-19 vaccination administered since the start of the vaccination roll-out on 8 December 2020, and who were aged 12-29 years on the date of their last vaccination. There were insufficient numbers and follow-up to study vaccination in children below 12 years of age. In a separate analysis, we also included all unvaccinated people who died within 12 weeks of having received a positive SARS-CoV-2 test on or after 9 September 2020 (when mass testing became available) and who were aged 12-29 years on the date of positive test.

### Exposure and outcomes

The main exposure was time since COVID-19 vaccination, with only the last vaccination received before death being included. The second exposure was time since the date of a SARS-CoV-2 positive test. The date of positive test is defined as the start of the last episode of COVID-19 for each individual, where a new episode is a positive result more than 120 days after the start of any previous COVID-19 infection episode. Only individuals where the SARS-CoV-2 positive tests occurred after the introduction of mass testing on 9 September 2020 are included in the analysis.

Two outcomes, occurring within 12 weeks of vaccination or positive test, were analysed: firstly, cardiac death (ICD-10 code I30-I52 mentioned on the death certificate); secondly, deaths due to all causes. Only participants who experienced both the exposure and outcome of interest were included in the study population.

### Statistical analysis

We used a self-controlled case series design, which compares the incidence of the outcome in a risk period to a baseline period to assess whether there is a change in the risk of death soon after vaccination compared to later after vaccination.

Follow up started on the day of last vaccination received and participants were not censored if a death occurred but were followed for 12 weeks after vaccination or until the end of study if sooner [11]. The follow up time was restricted to 12 weeks to minimise the impact of registration delay, where deaths that occurred in later calendar weeks were less likely to have been registered.

The follow up period was divided into weeks: the risk period was defined as the first 6 weeks after vaccination, the baseline period as the following 6 weeks. Myocarditis tends to appear very soon after vaccination, with evidence of the median time from vaccination to symptom onset of 2 days [12]. However, we used the first 6 weeks after vaccination as the risk period to ensure that all deaths resulting from myocarditis would be captured.

The exposure was the week since vaccination for each of the first 6 weeks or the whole risk period, and the baseline period. In a self-controlled case study, all fixed-time covariates are controlled for, as each participant acts as their own control, therefore no covariates were included in the main analysis. However, the calendar day of the start of each week was included as a time varying covariate in a sensitivity analysis.

The self-controlled case series models were fitted using a conditional logistic regression model on a person-week level dataset, with an individual effect [13]. Relative incidence of cardiac or all-cause deaths in risk periods relative to baseline periods, and their 95% confidence intervals were estimated using each model.

Similar analyses were conducted focusing on unvaccinated individuals who died within 12 weeks of a positive test for SARS-CoV-2.

All analyses were conducted using R 3.5

### Subgroup analyses

Analyses were stratified by sex, age groups (12-17, 18-24, 25-29), vaccine type (mRNA, not mRNA) and last vaccination dose received (first, second or third/booster).

### Sensitivity tests

Because of registration delays, deaths that occurred in later calendar weeks are less likely to have been registered than those which occurred in earlier calendar weeks (Supplementary Fig. 1). This affects the later follow up weeks after vaccination more than earlier, as a greater proportion of the later follow up weeks are later calendar weeks, and therefore a higher proportion of the deaths that occurred later after vaccination rather than earlier will be missing from our dataset (Supplementary Fig. 2). This is particularly apparent for young people, where a higher proportion of deaths will go to inquest and therefore take a longer time to be registered [13]. To mitigate this issue of registration delay, we restricted follow up time to 12 weeks after vaccination and conducted sensitivity analyses with different number of weeks of for the baseline period. Sensitivity tests were also carried out with different lengths of risk period to assess the impact of changing the definition of risk and control periods.

A sensitivity analysis was run where the calendar day of the start of each week since vaccination was included in the model as a spline, which will reflect the impact of seasonal mortality and changing SARS-CoV-2 infection rates.

### Patient and public involvement

No patient or member of the public was involved in this study

## Results

Table 1 shows the characteristics of the study population for the analyses of cardiac and all-cause deaths for the main analysis of deaths after vaccination for 12-29 year olds. Of the 585 deaths which occurred within 12 weeks of receiving a COVID-19 vaccination, 105 were due to a cardiac event. Of those who died, 54 (9.3%) were aged 12 to 17 years, 262 (44.8%) 18 to 24 years and 269 (46.0%) 25 to 29 years; 352 (61.0%) were male.

**Table 1.** Characteristics of the study population, young people aged 12-29, resident in England, who died within 12 weeks of receiving a COVID-19 vaccination, for deaths registered by 16 February that occurred by 2 February. Figures are number (%) unless stated otherwise.

| Characteristic | Level | All-cause deaths | Cardiac-related deaths |
| --- | --- | --- | --- |
| Age group | 12-17 | 54 ( 9.23) | 9 ( 8.57) |
|  | 18-24 | 262 (44.79) | 43 (40.95) |
|  | 25-29 | 269 (45.98) | 53 (50.48) |
| Calendar date of vaccination | (median) | 20/05/2021 | 07/06/2021 |
| Time since vaccination | 6 weeks or less | 293 (50.09) | 51 (48.57) |
|  | 7 to 12 weeks | 292 (49.91) | 54 (51.43) |
| Sex | Male | 352 (60.17) | 64 (60.95) |
|  | Female | 233 (39.83) | 41 (39.05) |
| Vaccination | First | 315 (53.85) | 49 (46.67) |
|  | Second | 208 (35.56) | 39 (37.14) |
|  | Third | 62 (10.60) | 17 (16.19) |

Of the 212 deaths which occurred within 12 weeks of a positive test for SARS-CoV-2 taken from 9 September onwards, 22 were due to a cardiac event.

### Relative incidence of death after COVID-19 vaccination

Figure 1 shows the relative incidence of cardiac deaths and all-cause deaths in each of the first 6 weeks after vaccination compared to the control period, and for the first six weeks as a whole compared to the baseline period.

**Fig 1.**
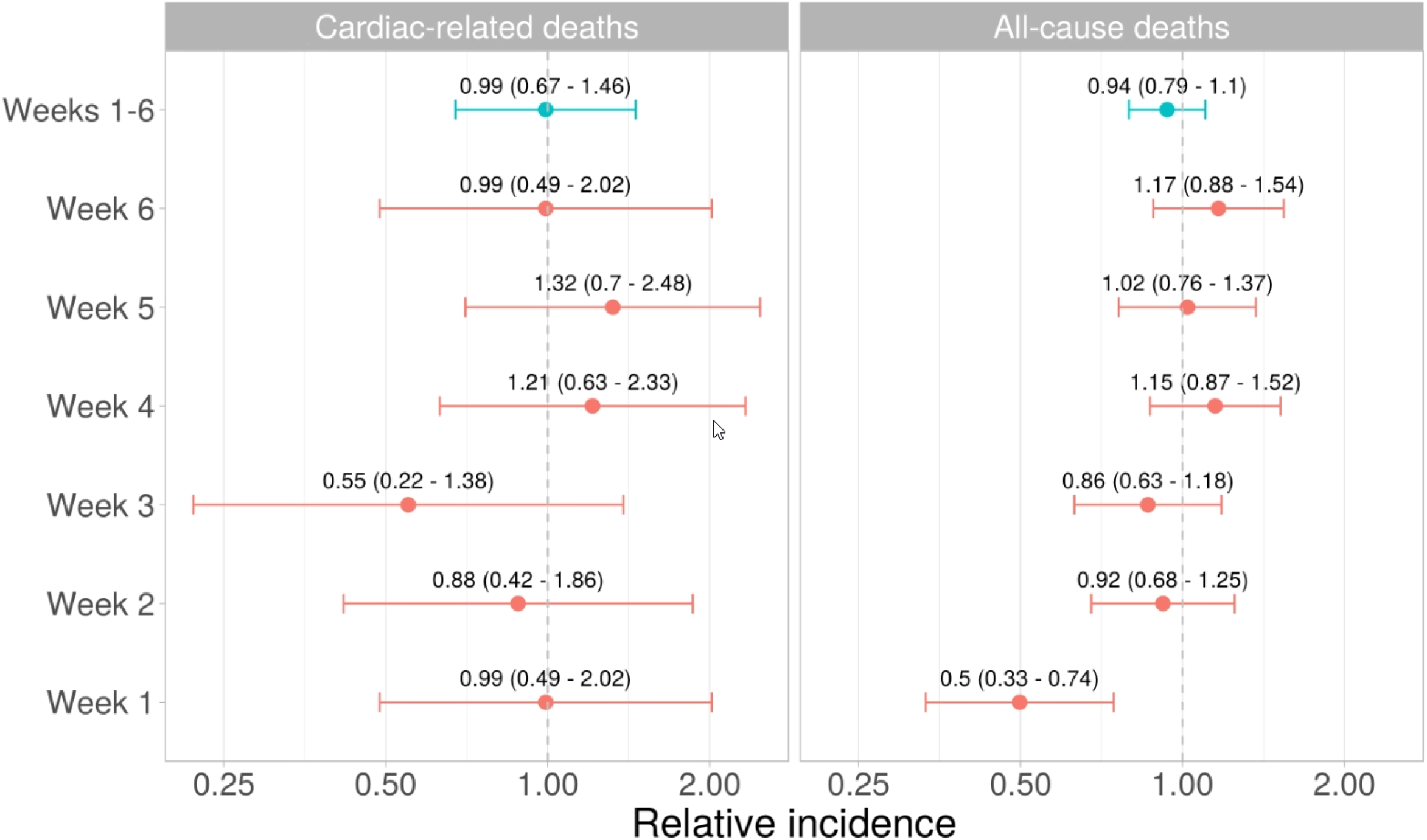
Relative incidence (95% confidence intervals) of cardiac death and all-cause death in the each of the 6 weeks in the risk period after the last vaccination received and in the risk period as a whole, compared to the baseline period.

There was no evidence of a change in the risk of cardiac death in any of the first 6 weeks in the risk period after vaccination or in the risk period as a whole (relative incidence 0.99, 95% confidence interval 0.67 to 1.46).

There was no evidence of an elevated risk of all cause death in any of the first 6 weeks in the risk period after vaccination and no change in the risk of all cause death in the risk period as a whole (0.94, 0.79 to 1.10). A decrease in the risk of death for all causes was observed in the first week after vaccination (0.50, 0.33 to 0.74).

Figure 2 shows the relative incidence of death in the risk period (weeks 1 to 6) after vaccination compared to the baseline period, stratified by sex, age group, vaccine type and last dose received. For all breakdowns, there was no evidence of a change in the risk of death for either cardiac or all-cause deaths in the first six weeks after vaccination.

**Fig 2.**
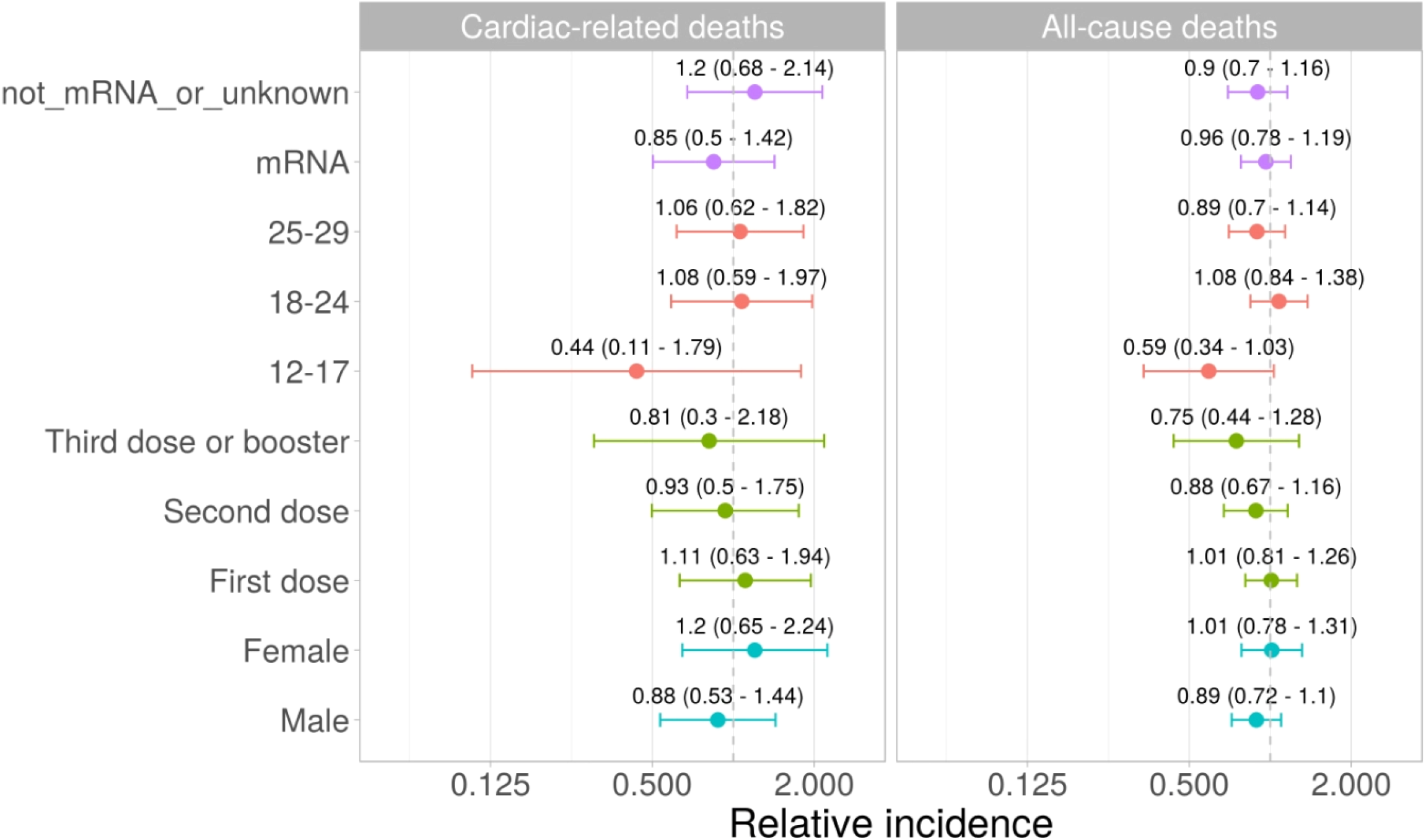
Relative incidence (95% confidence intervals) of cardiac and all-cause death in the risk period after the last vaccination received, compared to the baseline period.

### Relative incidence of death after SARS-CoV-2 infection

Figure 3 shows that there was an increase in the risk of cardiac death in the overall risk period after SARS-CoV-2 infection (6.00; 95% CI: 1.77 to 20.37) with the largest increase in risk during the first week (15.00; 3.96 to 56.88). Increases in the risk of all-cause mortality were also observed in the whole risk period after SARS-CoV-2 infection (4.42; 3.10 to 6.32), with the largest increase in risk occurring in the first week (7.56; 4.93 to 11.58). Characteristics for the study population used in this analysis are shown in Supplementary Table 1.

**Fig 3.**
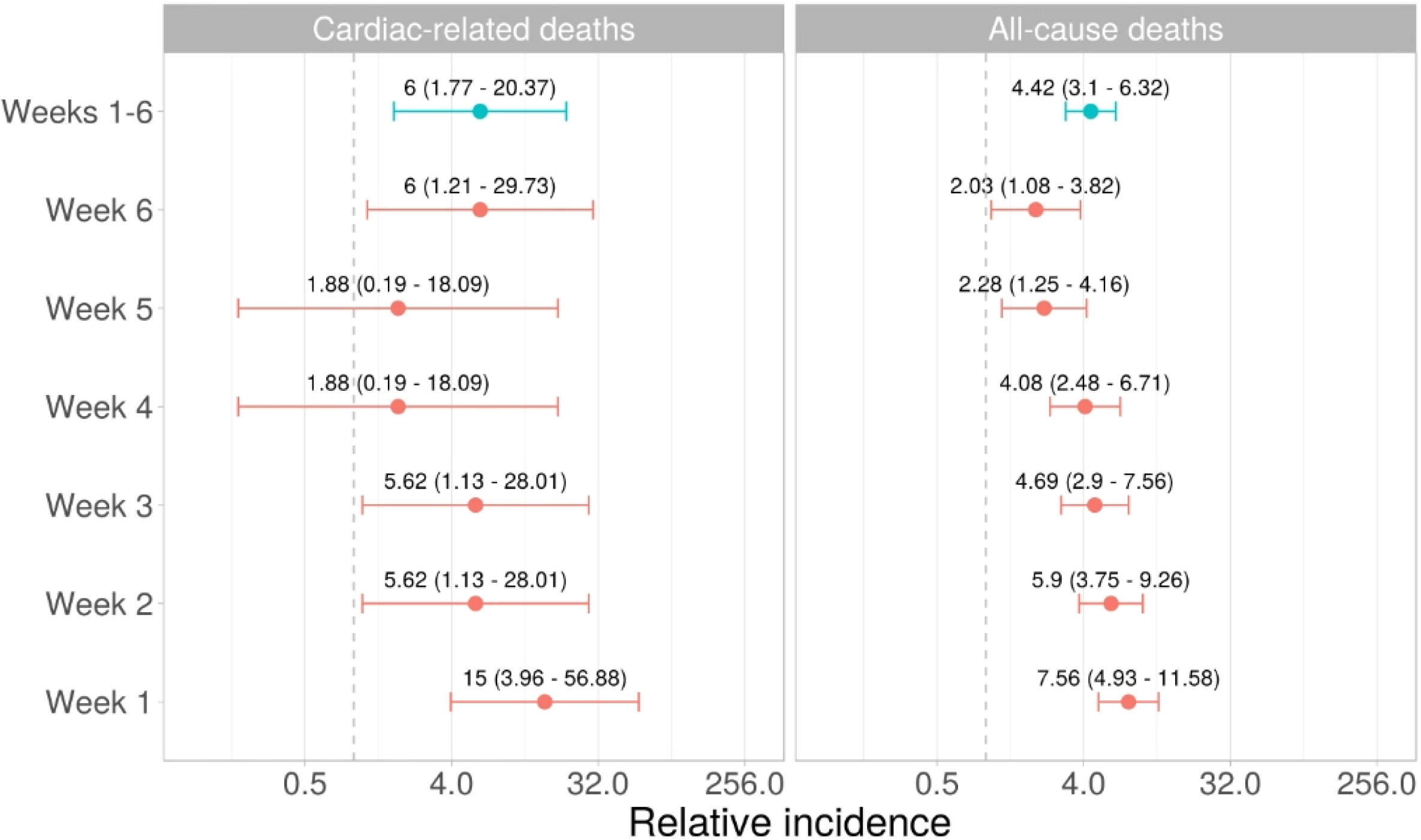
Relative incidence (95% confidence intervals) of cardiac and all-cause death in the risk period after the most recent SARS-CoV-2 infection, compared to the baseline period.

### Sensitivity tests

Supplementary Figure 2 shows the number of deaths each week since vaccination. Later weeks since vaccination are more likely to be affected by the decreased likelihood of a death being registered due to registration delay. There is a drop off around 12 weeks, where we set the cut-off of the control period in our analysis.

Including the calendar time of each week as a covariate in the conditional logistic regression models did not produce significantly different results (Supplementary Figure 3). Changing the number of weeks after vaccination that are included in the risk period, while keeping the follow up at 12 weeks, did not result in a change in the relative incidence for any risk period length for cardiac deaths (Supplementary Figure 4). However, for all-cause deaths, results were unstable when selecting a small number of weeks (2 or 3).

Because of registration delays, increasing the follow up time also increases the relative incidence rate for the 6-week risk period (Supplementary Figure 5). The estimates of relative incidence become statistically significant for cardiac death if follow up is extended to 18 weeks or beyond; for all-cause death, it becomes statistically significant if follow up is extended to 16 weeks or beyond.

## Discussion

### Main findings

Using a self-controlled case series approach, we found no evidence of an increased risk in cardiac death following a COVID-19 vaccination. Whilst the risk of all-cause death in the first week after vaccination was lower than in the baseline, it was not different from the baseline in each of weeks 2 to 6 after vaccination. We also found no evidence of increased mortality after vaccination for either cardiac or all cause in any subgroup. By contrast, we observed an increase in the risk of cardiac deaths and all-cause deaths after SARS-CoV-2 infection.

### Comparison with other studies

Several studies have highlighted the association between COVID-19 vaccination and the risk of myocarditis and other cardiac events. Vaccination with mRNA vaccines is associated with an increased risk of myocarditis or myopericarditis, especially in young people, with data from the US [12], Denmark [6], and England [14]. We found no evidence of increased risk of death due to cardiac events, which suggest that the cases of myocarditis or myopericarditis due to the COVID-19 vaccination are unlikely to be fatal. However, whilst these cases of myocarditis or myopericarditis may not result in death, they may lead to the development of longer term cardiac complications.

Our results on the raised risk of deaths due to cardiac events following SARS-CoV-2 infection are also consistent with evidence from Sweden showing elevated risk of myocardial infarction and ischaemic stroke following COVID-19 [8].

This lower risk of all-cause death in the first week following vaccination provides evidence of a ‘healthy vaccinee effect’, where people who are unwell are more likely to delay vaccination until recovered, therefore the health of people who have recently received a vaccination is generally better than that of those who are eligible. This healthy vaccinee effect is commonly observed in studies estimating vaccine effectiveness [15].

### Strengths and limitations

Our study has several strengths. First, we used death registration records for the whole of England, linked to all vaccination records, including those which are not available in the main NIMS COVID-19 vaccination data, because the people died shortly after vaccination. As a result, we had information on the cause of death based on medical certificate of cause of death. To limit further the risk of outcome misclassification, we not only examined deaths due to cardiac events but also all-cause mortality. Second, using a self-controlled case series, our estimates account for time-invariant confounding, which is crucial because young people who were clinically extremely vulnerable were prioritised for vaccination. Comparing the risk of deaths between vaccinated and unvaccinated individuals would have been challenging in the absence of detailed information on pre-existing health status.

The main limitation of our study is the delay in death registration. Not all deaths that occurred in the period have already been registered and registration delays can be substantial for young people, as deaths are more likely to be investigated by a coroner than for older adults. Registration delays should affect more deaths that occurred later in the year, and therefore deaths that occurred further from vaccination status, leading to overestimating the incidence case ratios. To minimise the impact of registration delays, we restricted the follow up period to 12 weeks and conducted some sensitivity analyses. The SCCS relies on several assumptions. First, the assumption that deaths are rate. Among 12-29 year olds, the mortality rate was very low and unlikely to bias in the estimates of the relative incidences. Another limitation is that we could not control for SARS-CoV-2 infection [16], despite the infection being related to cardiovascular outcomes [8].

## Conclusion

Whilst COVID-19 vaccination has been linked to an increased risk of myocarditis and other cardiac events in young people, our study shows that there is no evidence of increased risk of death due to cardiac events, which suggest that cases of myocarditis or myopericarditis due to the COVID-19 vaccination are unlikely to be fatal. This provides reassurance that the benefits of COVID-19 vaccines outweigh the risks even in young people.

## Data Availability

Data used in this study will be made available in the ONS Secure Research Service

## Acknowledgements

We thank James Doidge, Nick Andrews and Chris Robertson for useful discussions. KK is supported by the National Institute for Health Research (NIHR) Applied Research Collaboration East Midlands (ARC EM) and the NIHR Leicester Biomedical Research Centre (BRC).

## Ethical approval

Ethical approval was obtained from the National Statistician’s Data Ethics Advisory Committee (NSDEC(20)12).

## Data availability

Data used in this study will be made available in the ONS Secure Research Service

## Supplementary Information

**Supplementary Figure 1.**
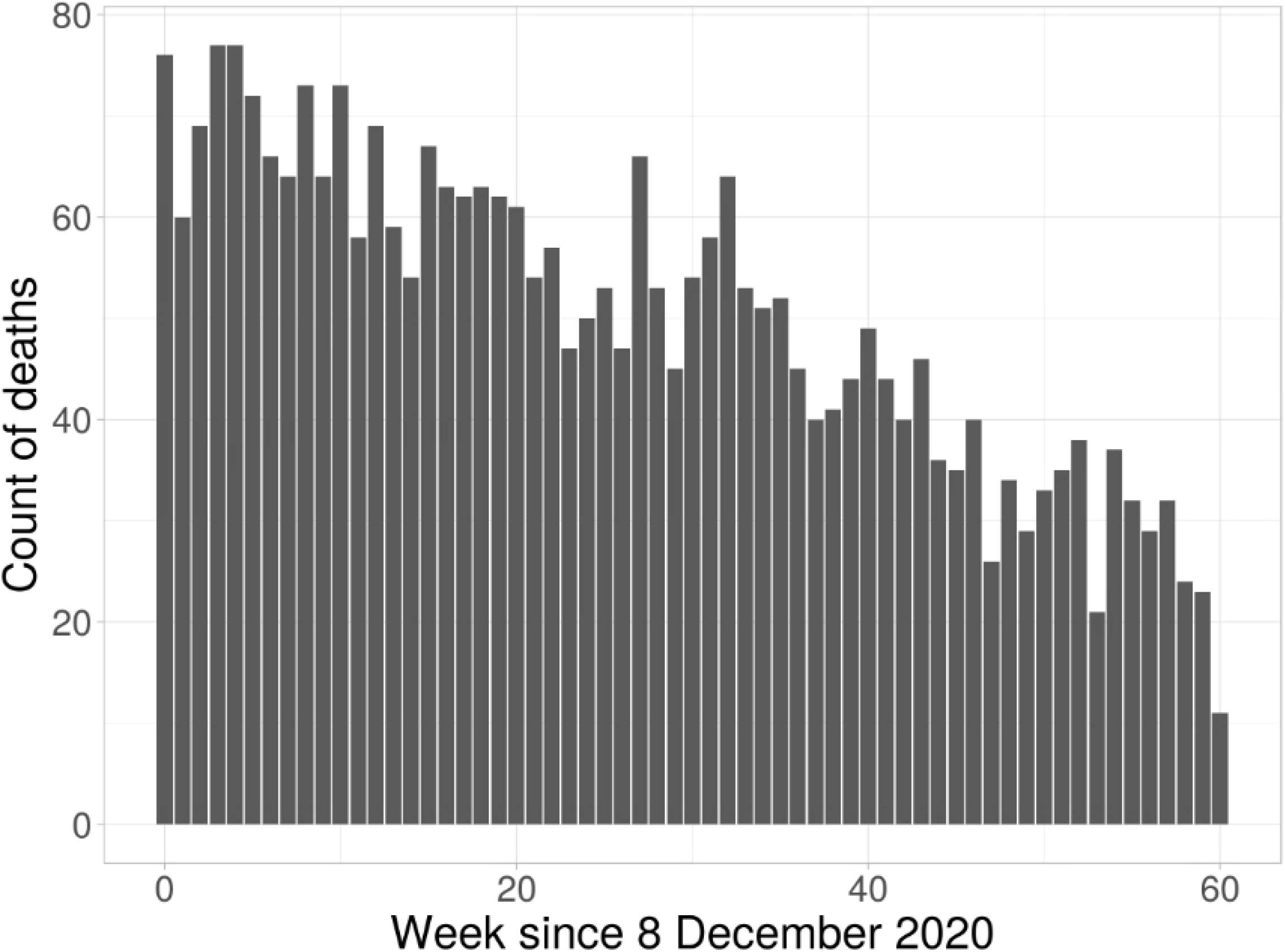
Number of deaths that occurred each week for weeks since December 8 2020, and were registered by 13 February 2022, young people aged 12-29 on date of death. Data is for all deaths, not just those of vaccinated people.

**Supplementary Figure 2.**
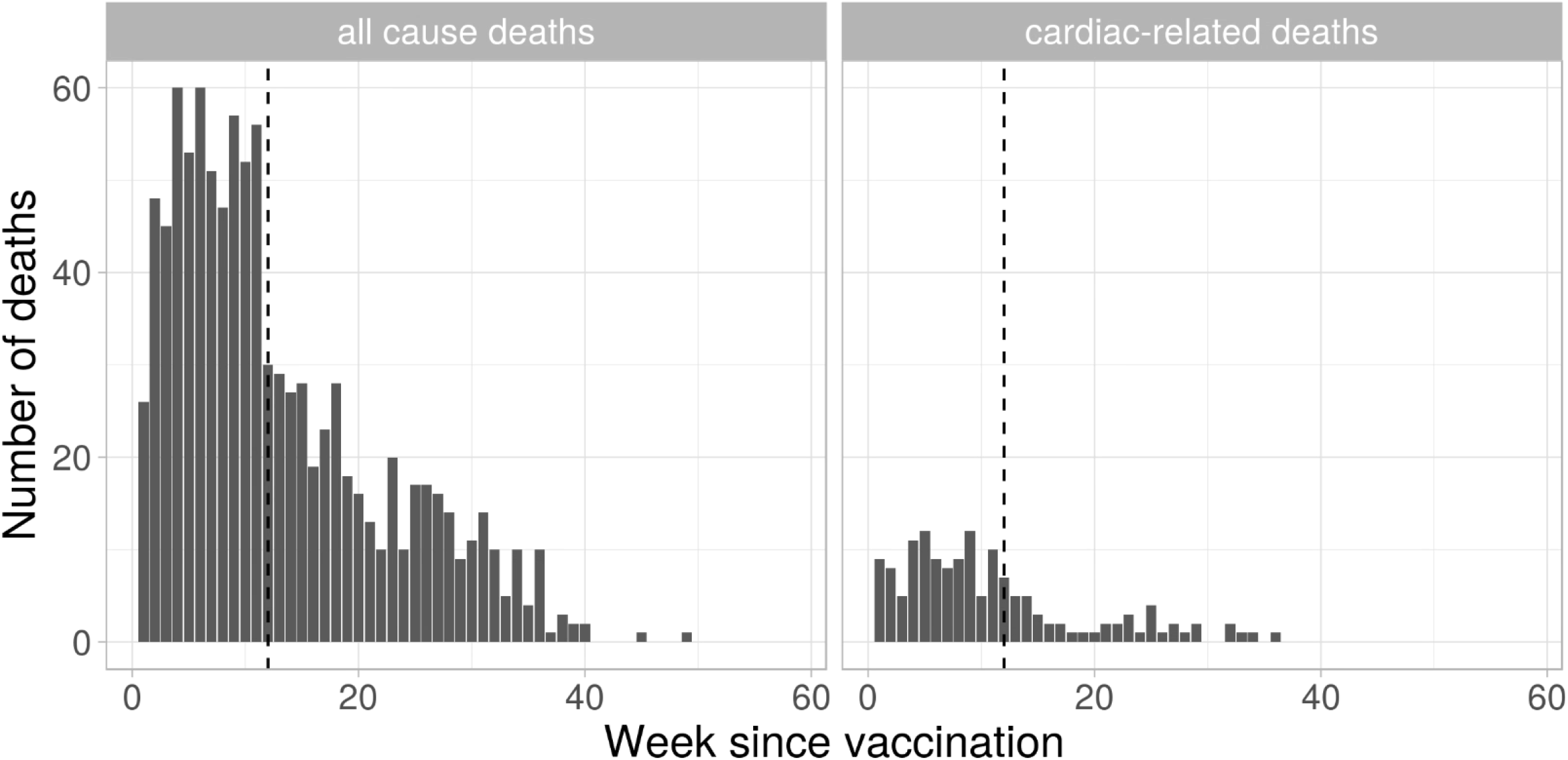
Number of deaths each week since vaccination. Week 12 is indicated by the dotted line.

**Supplementary Figure 3.**
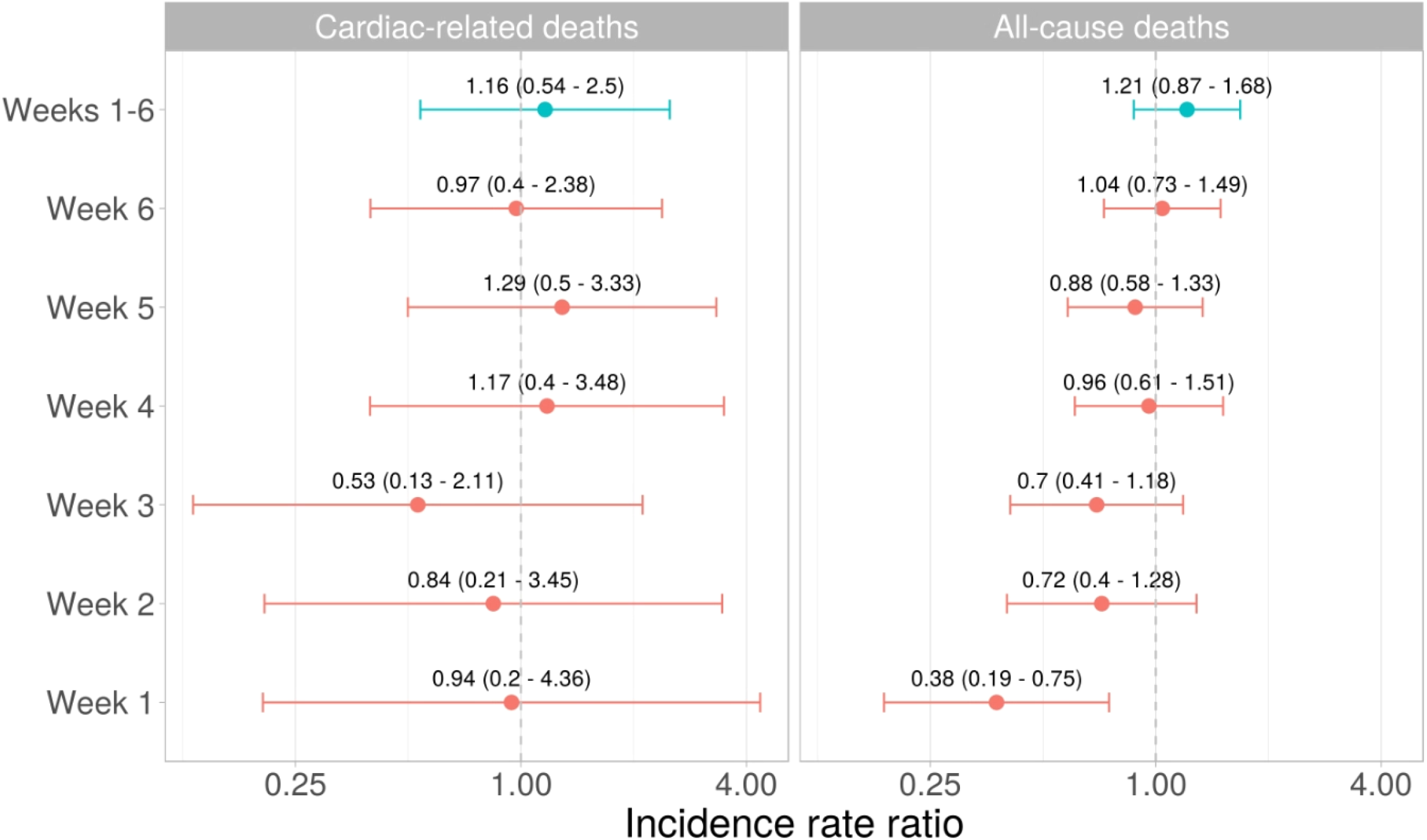
Relative incidence of cardiac and all-cause deaths in the risk period compared to the control period, with calendar time as a covariate.

**Supplementary Figure 4.**
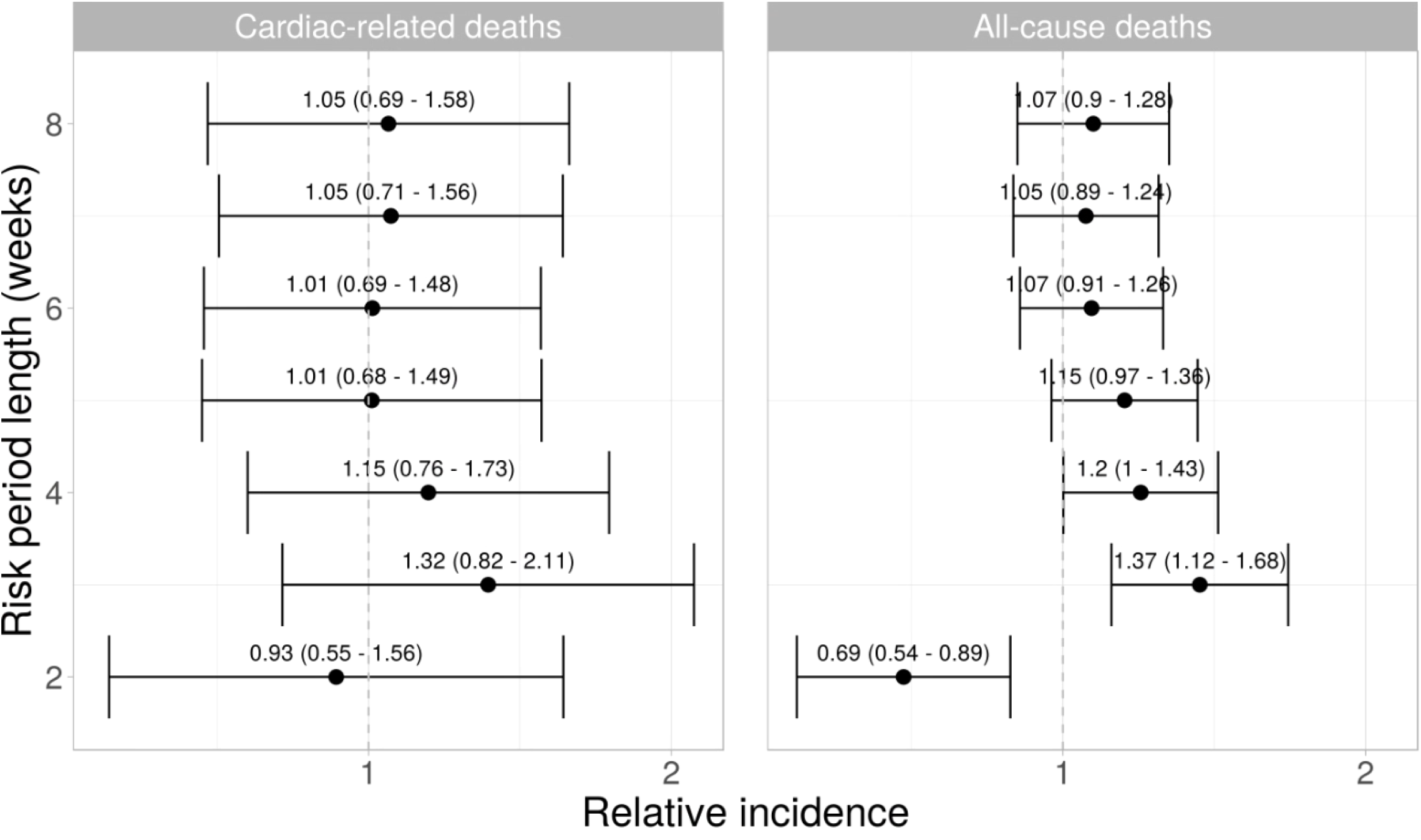
Relative incidence of cardiac and all-cause deaths in the risk period compared to the baseline period, for different lengths of risk period.

**Supplementary Figure 5.**
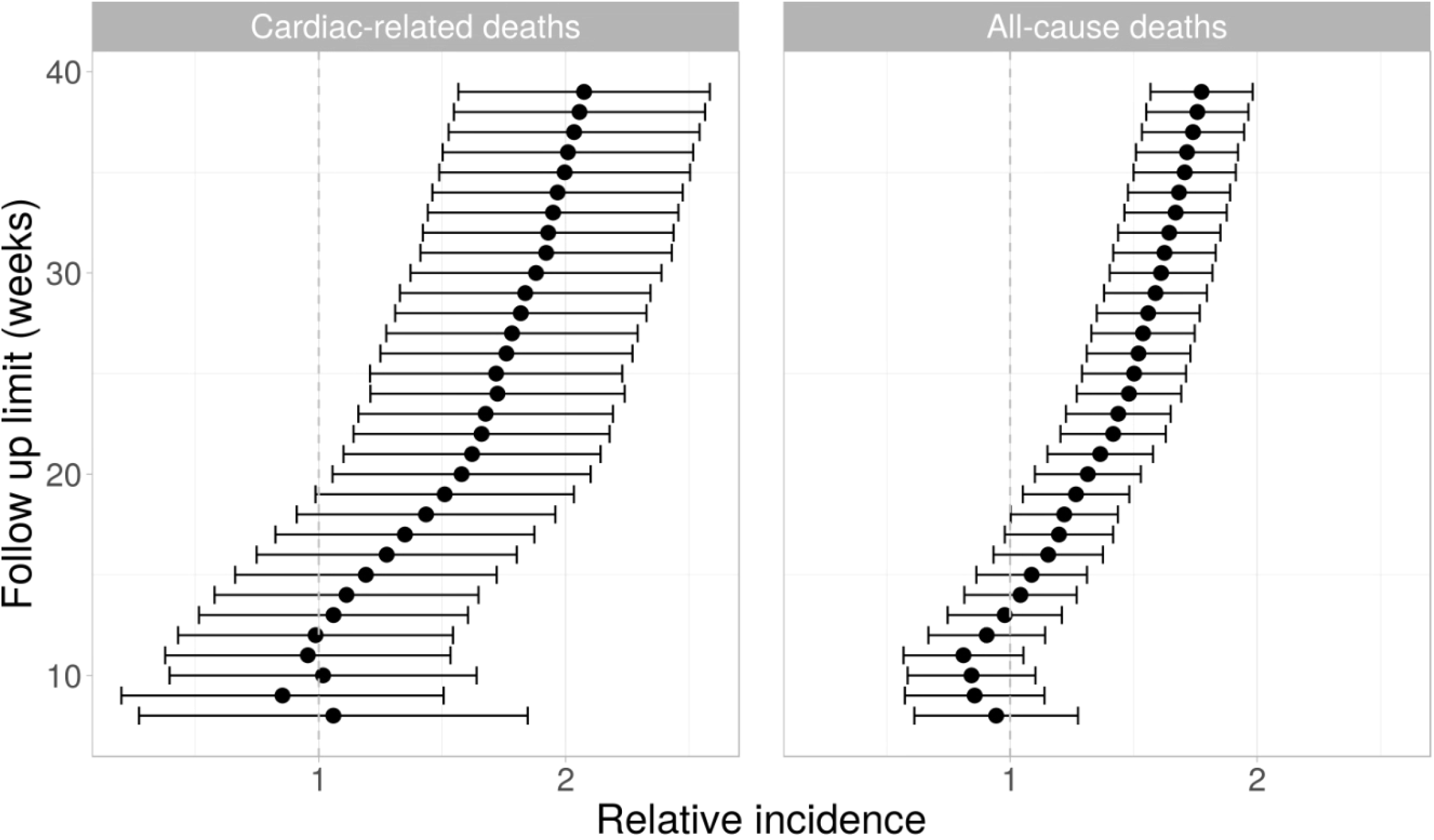
Relative incidence of cardiac and all-cause deaths in the risk period compared to the baseline period, for different lengths of follow up.

**Supplementary Table 1.** Characteristics of the study population for analysis of death after SARS-CoV-2 infection, unvaccinated young people aged 12-29, resident in England, who died within 12 weeks of the start of a SARS-CoV-2 infection, for deaths registered by 16 February that occurred by 2 February. Figures are number (%) unless stated otherwise.

| Characteristic | Level | All-cause deaths | Cardiac-related deaths |
| --- | --- | --- | --- |
| Age group | 12-17 | 21 ( 9.91) | 4 (18.18) |
|  | 18-24 | 84 (39.62) | 8 (36.36) |
|  | 25-29 | 107 (50.47) | 10 (45.45) |
| Calendar date of infection | (median) | 08/02/2021 | 05/02/2021 |
| Time since infection | 6 weeks or less | 175 (82.55) | 19 (86.36) |
|  | 7 to 12 weeks | 37 (17.45) | 3 (13.64) |
| Sex | Male | 130 (61.32) | 17 (77.27) |
|  | Female | 82 (38.68) | 5 (22.73) |

